# Detecting neurodegenerative changes in glaucoma using deep mean kurtosis-curve–corrected tractometry

**DOI:** 10.1101/2025.06.05.25329075

**Authors:** Loxlan W. Kasa, William Schierding, Eryn Kwon, Samantha Holdsworth, Helen V Danesh-Meyer

## Abstract

Glaucoma is increasingly recognized as a neurodegenerative condition involving both retinal and central nervous system structures. Here, we present an integrated framework that combines MK-Curve-corrected diffusion kurtosis imaging (DKI), tractometry, and deep autoencoder-based normative modeling to detect localized white matter abnormalities associated with glaucoma. Using UK Biobank diffusion MRI data, we show that MK-Curve approach corrects anatomically implausible values and improves the reliability of DKI metrics – particularly mean (MK), radial (RK), and axial kurtosis (AK) – in regions of complex fiber architecture. Tractometry revealed reduced MK in glaucoma patients along the optic radiation, inferior longitudinal fasciculus, and inferior fronto-occipital fasciculus, but not in a non-visual control tract, supporting disease specificity. These abnormalities were spatially localized, with significant changes observed at multiple points along the tracts. MK demonstrated greater sensitivity than MD and exhibited altered distributional features, reflecting microstructural heterogeneity not captured by standard metrics. Node-wise MK values in the right optic radiation showed weak but significant correlations with retinal OCT measures (ganglion cell layer and retinal nerve fiber layer thickness), reinforcing the biological relevance of these findings. Deep autoencoder-based modeling further enabled subject-level anomaly detection that aligned spatially with group-level changes and outperformed traditional approaches. Together, our results highlight the potential of advanced diffusion modeling and deep learning for sensitive, individualized detection of glaucomatous neurodegeneration and support their integration into future multimodal imaging pipelines in neuro-ophthalmology.

## 1. Introduction

Glaucoma is a leading cause of irreversible blindness worldwide and is characterized by progressive degeneration of retinal ganglion cells and the optic nerve head, leading to visual field loss (Kang & Wan, 2022; Weinreb et al., 2014). Although traditionally viewed as an eye disease, glaucoma is increasingly recognized as a neurodegenerative disorder that affects the entire visual pathway, and even non-visual brain regions – through trans-synaptic degeneration (Y. Sun et al., 2019; Torres & Hatanaka, 2019). Early detection remains a critical challenge due to the disease’s heterogeneous and often asymptomatic progression, contributing to its status as one of the most underdiagnosed neuro-ophthalmic conditions (Nayak et al., 2011; Wong et al., 2004). These demonstrate the need for advanced, sensitive neuroimaging techniques capable of detecting early and spatially distributed brain changes (Bourne & Khatib, 2021; Sharma et al., 2008).

Diffusion-weighted MRI (dMRI) enables non-invasive characterization of brain tissue microstructure by quantifying the movement of water molecules. Diffusion tensor imaging (DTI), a widely used dMRI technique, has been utilized to assess glaucoma-related changes along the visual pathway using parameters such as fractional anisotropy (FA) and mean diffusivity (MD). While DTI-based studies have revealed abnormalities beyond the optic nerve including the lateral geniculate nucleus (LGN), optic radiations (OR), and visual cortex, DTI is limited by its assumption of Gaussian diffusion, which fails to capture microstructural complexity in regions with crossing fibers or heterogeneous tissue composition (Basser & Pierpaoli, 1996; Jensen et al., 2005; J.-D. Tournier et al., 2011).

Diffusion kurtosis imaging (DKI) extends DTI by modeling non-Gaussian diffusion, providing additional markers such as mean kurtosis (MK), axial kurtosis (AK), and radial kurtosis (RK), which are sensitive to tissue complexity and glial proliferation (Jensen & Helpern, 2010; Steven et al., 2014). Initial DKI studies in glaucoma have reported MK reductions in visual pathway structures, with evidence of widespread alterations using tract-based spatial statistics (TBSS) (Xu et al., 2018; Nucci et al., 2020). However, voxel-wise methods like TBSS require accurate registration across individuals and are limited in their ability to capture tract-specific pathology, especially when dealing with a clinical cohort (Tsang et al., 2010; Yeatman et al., 2012).

To overcome these limitations, recent studies in glaucoma have utilized tractometry, an approach that quantifies tissue properties along fiber tracts reconstructed from tractography (Kasa et al., 2025). Kruper et al. (2024) applied automated fiber quantification (AFQ) framework (Yeatman et al., 2012) for DKI-based tractometry in the optic radiations using data from the UK Biobank (UKBB) (Sudlow et al., 2015), combining it with convolutional neural networks (CNNs) for glaucoma classification. Their findings demonstrated that localized abnormalities in DKI metrics are predictive of disease, underscoring the potential of DKI to detect microstructural damage in glaucoma. However, while DKI provides more biologically sensitive measures than DTI, it is also highly susceptible to noise, often leading to implausible kurtosis estimates that limits its reliability in large-scale studies; the MK-Curve correction method addresses this issue by effectively recovering voxel-wise implausible values without the need for smoothing (Christiaanse et al., 2023; Zhang et al., 2019). Furthermore, there is growing interest in shifting from group-level analyses to individualized anomaly detection to enable potential clinical translation. Deep learning algorithms, such as autoencoders (AE), are increasingly used to identify subtle deviations from normative patterns at the single-subject level (Chamberland et al., 2021), enhancing clinical applicability.

In this work, we sought to evaluate the utility of MK-Curve-corrected DKI tractometry combined with deep autoencoders for detecting microstructural abnormalities in glaucoma, leveraging the large-scale UKBB diffusion MRI dataset to reduce sample bias and enhance generalizability. To extract tissue properties along white matter (WM) bundles we used BundleSeg (St-Onge et al., 2023), a robust bundle segmentation method. We hypothesize that integrating MK-Curve-corrected diffusion kurtosis imaging with deep autoencoder-based anomaly detection will enhance the sensitivity and spatial specificity of detecting microstructural abnormalities in glaucoma, particularly within the optic radiation (OR) and associated WM tracts, while enabling subject-level anomaly detection.

**Figure 1.**
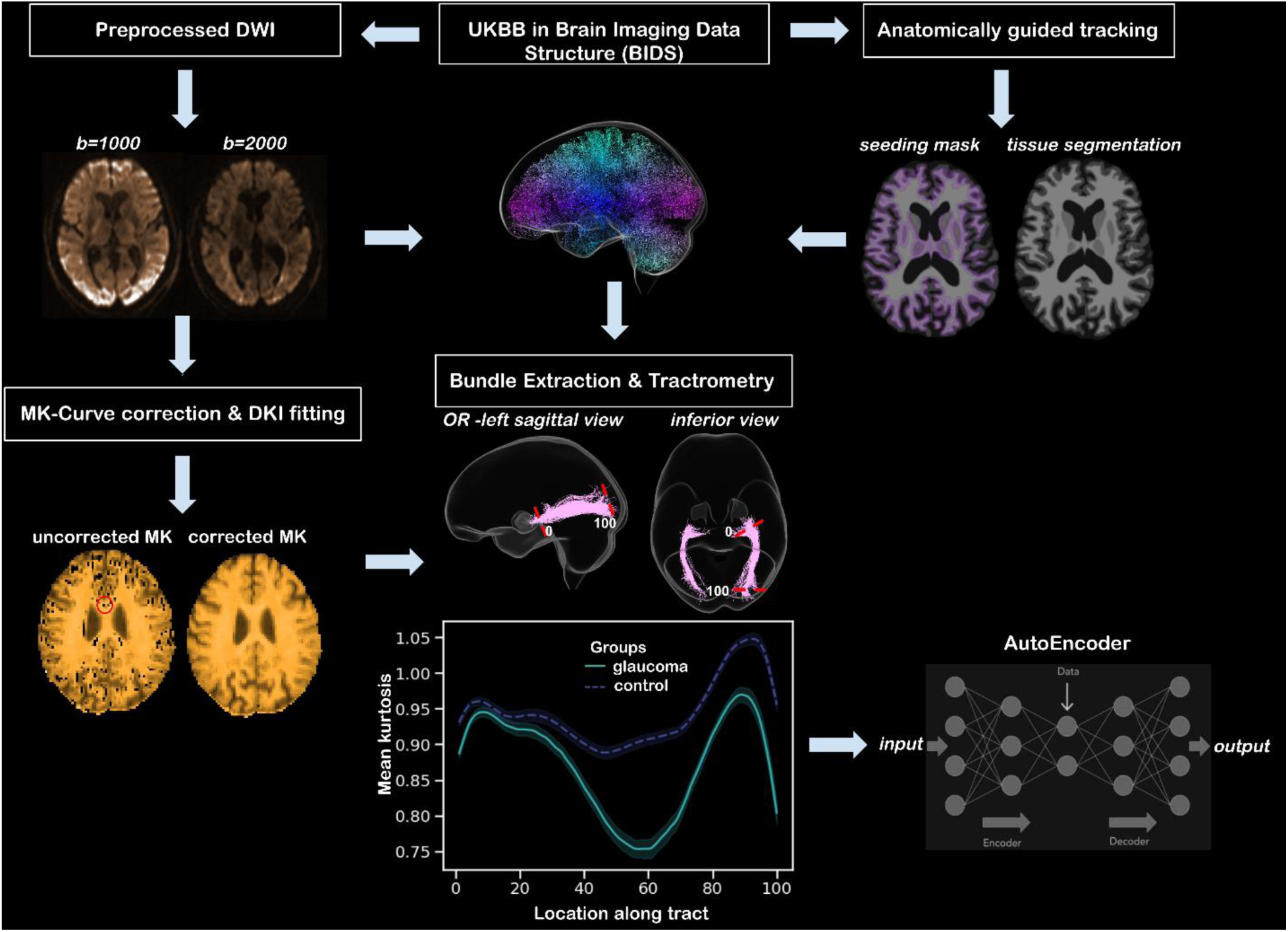
Overview of the analysis pipeline. UK Biobank (UKBB) diffusion and structural MRI data were first converted to Brain Imaging Data Structure (BIDS) format. Preprocessed DWI volumes were then corrected using the MK-Curve approach before the computation of diffusion kurtosis imaging (DKI) parametric maps - mean (MK), radial (RK) and axial kurtosis (AK). Anatomically constrained tractography was used to generate whole-brain streamlines utilizing T1-weighted images and preprocessed DWIs, followed by bundle extraction and processing. Tractometry was performed along each bundle using the MK-Curve corrected DKI maps. Finally, a deep autoencoder was trained on the resulting tract profiles to identify white matter anomalies in glaucoma patients.

## 2. Materials and Methods

### 2.1 Cohort derivation and characteristics

Data for this study were sourced from the UKBB health research program (Sudlow et al., 2015). We obtained de-identified MRI and health data from the UKBB repository. The study was approved by the UKBB Ethics Committee (11/NW/0382) and conducted under UKBB application number 76765. Participants provided informed consent during a detailed recruitment visit, which included broad consent for using their anonymized data in diverse health research. (Sudlow et al., 2015).

### 2.1.1 Glaucoma definition

Glaucoma cases within the UKBB cohort were identified using three data sources: self-reported diagnoses, hospital inpatient records, and primary care data. A participant was classified as a glaucoma case if they had a positive record in any of these sources, corresponding to UKBB fields 6148, 20002, 41271 (ICD9 365.1, 365.8, 365.9), 41270 responses (ICD10 H40.1, H40.8, or H40.9), or 42040 (GP Read2/Read3 mapping to the ICD9 and ICD10 codes). To identify individuals with potentially more advanced disease, a subset of glaucoma cases was defined as “diagnosed-and-treated” based on evidence of treatment via surgical intervention and/or medication (UKBB fields 5326, 5327, 20003, 20004, 41272, and 41273). Controls were defined as UKBB participants with no history of glaucoma, including those with subtypes not considered in the case definition (*e.g.*, angle-closure glaucoma, glaucoma suspects). To address case-control imbalance and control for confounding by age and sex – both known to influence glaucoma risk and MRI outcomes, controls were matched to cases using the MatchIt package in R (v4.2.1), providing exact matching on age and sex across all cases.

### 2.1.2 Sample selection

The UKBB cohort was divided into Glaucoma and Control groups based on glaucoma status as defined above in individuals with available MRI data: 1) All diagnosed glaucoma cases (n=1,465); 2) The control cohort was selected from individuals with MRI data available (n=41,736). Of the 1,465 glaucoma cases, only 717 had evidence of treatment. Of those, only 404 had MRI data across all necessary MRI scans. The 404 glaucoma cases were age and sex matched with controls with valid MRI scans (n=419 controls).

### 2.2 Image acquisition and processing

The preprocessed UKBB dMRI dataset was used for this study. Here we have provided a brief overview of the acquisition protocol, and the details have been reported elsewhere (Miller et al. 2016). Imaging was done on a 3T Siemens Skyra scanner with a 32-channel radiofrequency receiver head coil, using a standardized acquisition protocol. The scanning protocol included the acquisition of structural images using a magnetization-prepared rapid acquisition with gradient echo (MPRAGE) sequence, with inversion and repetition times optimized for maximal contrast, FOV = 208×256 mm^2^ and 1 mm isotropic voxel size. Diffusion MRI acquisition parameters include TR/TE = 3600/92 ms, FOV = 104 × 104 mm^2^, 2 mm isotropic voxel size, a multiband echo-planar imaging (EPI) sequence with an acceleration factor=3. A total of 100 diffusion weighted volumes were acquired using distinct anterior-to-posterior phase encoding directions at b=1000 s/mm^2^ and b=2000 s/mm^2^, with 5 non-diffusion weighted volumes at b=0 s/mm^2^. In addition, extra 3 volumes at b=0 s/mm^2^ were acquired with posterior-to-anterior phase encoding direction and subsequently used for EPI distortion correction. The detail of the diffusion MRI preprocessing pipeline has also been discussed elsewhere (Alfaro-Almagro et al. 2018), and here we have provided only a short overview. In brief, dMRI preprocessing included head motion and eddy currents correction using the FSL “eddy” software, with correction of outlier slices followed by gradient distortion correction.

### 2.3 DKI quantitative map estimation and correction

The general DKI signal equation (below) is an expansion from the common DTI signal equation which allows DKI to estimate both DKI and DTI quantitative maps (Jensen et al., 2005; Wu & Cheung, 2010).

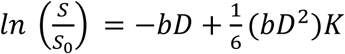

Where *S* is the signal measured at *b*≠ 0 (diffusion weighting) along a specific direction, *S_0_* is the non-diffusion-weighted signal measured at *b*= 0, *D* is the diffusion tensor and *K* the fourth-order kurtosis tensor. DKI attempts to characterize the degree to which diffusion deviates from Gaussian behavior, for which kurtosis (*K*) is zero. Kurtosis is a dimensionless statistic and mathematically can take values from positive where the displacement distribution curve has a more sharply peaked profile or negative kurtosis with the distribution least peaked. Although mathematically kurtosis can have negative values, biological tissues have been shown to only exhibit positive kurtosis (Jensen et al., 2005; Tabesh et al., 2011). To estimate DKI quantitative maps will require determining 22 unique parameters (*S_0_*, six parameters of the symmetric tensor *D*, and 15 parameters of the symmetric tensor *K*). The 22 free parameters and the presence of a quadratic term linking the kurtosis and diffusion tensor results in the DKI model fitting to be sensitive to noise amplification (Christiaanse et al., 2023; Zhang et al., 2021). In this work, we used the MK-Curve method to correct any spurious kurtosis measurements (Zhang et al., 2019). The MK-Curve method uses synthetic b=0 images to model the relationship between b=0 intensity and mean kurtosis (MK), identifying and correcting implausible MK values using voxel-specific thresholds defined by MK=0 and MK=max (Zhang et al., 2019). This correction method has been seen to improve the reliability of DKI, particularly in regions prone to noise or artifacts (Christiaanse et al., 2023; Zhang et al., 2021). The UKBB DWI preprocessed data was further corrected with the MK-Curve correction method before calculating the DKI quantitative maps namely mean kurtosis (MK); radial kurtosis (RK); axial kurtosis (AK), and the common mean diffusivity (MD) from DTI.

### 2.4 MK-Curve corrected tractometry

We followed the Bundle Analytics (BUAN) framework (Chandio et al. 2020) to quantify the optic radiation (OR) and two adjacent networks (*i.e.*, inferior longitudinal fasciculus (ILF) and inferior fronto-occipital fasciculus (IFOF), which are involved in higher-order visual processing functions (Herbet et al., 2018; Sarubbo et al., 2013)). In addition, we included uncinate fasciculus (UF) as a control bundle. The main four steps of the BUAN framework were adopted into our workflow, which included (i) anatomically guided whole brain fiber tracking, (ii) bundle extraction using BundleSeg, (iii) tract-specific profile generation, and (iv) tractometry. Each step is further detailed in the following subsections.

#### 2.4.1 Anatomically constrained fiber tracking

For fiber tracking, we used the anatomically constrained tracking algorithm (ACT), which enhances the biological accuracy of diffusion MRI streamlines tractography reconstruction, by constraining the propagation and termination of streamlines based on tissue segmentation from a high-resolution, high-contrast anatomical image (*e.g.*, T1-weighted) (Smith et al. 2012). The ACT framework is implemented in MRtrix3 software (Tournier et al. 2019). The first step is to determine the response function. This was calculated from the UKBB preprocessed DWIs using *dwi2response* with the “dhollander” algorithm for multi-shell data (Dhollander et al. 2019). The response functions were estimated for individual DWIs which were then used for calculating the fiber orientation function (FOD) using *dwi2fod*. The FODs were separately calculated for the three tissue types (*i.e.*, WM, GM, and cerebrospinal fluid (CSF)) using the multi-shell multi-tissue constrained spherical deconvolution (CSD) approach (*msmt_csd*) (Jeurissen et al. 2014). To apply the ACT method, additional anatomical information is required to guide the tracking (Smith et al. 2012). All subjects’ T1 images were segmented into five tissue types (‘5TT’ – WM, subcortical gray matter (GM), GM, CSF and the optional pathological tissue which was excluded for this work). To minimize error in the segmentation due to presence of image noise and/or poor tissue contrast in the UKBB T1 scans, we applied the newly developed Hybrid Surface-Volume Segmentation (HSVS) technique (R. Smith et al., 2020). This step was accomplished using *5ttgen* with the HSVS algorithm (*5ttgen hsvs)* which took surface volume segmentation from FreeSurfer’s *recon-all* pipeline as input. We then generated a WM-GM interface mask by inputting the 5TT segmented anatomical image into the *5ttgmwmi* command to minimize tracking into the deep GM. This resulting WM-GM interface mask was then used as a seed point for tracking. For tractography, we employed the *tckgen* function with the *iFOD2* algorithm, which is adept at reconstructing fibers with complex configurations (J. D. Tournier et al., 2010) following the six criteria described by (R. E. Smith et al., 2012). To further guide fiber tracking, we used the following additional *tckgen* settings and inputs: step size = 0.8 mm, minimum length = 8 mm, maximum length = 250 mm, maximum number of streamlines = 10 million, and unidirectional tracking. The cutoff FOD amplitude was set to 0.06 during seeding, and streamlines were cropped at the GM-WM interface, with a more precise crop at the WM-GM interface. To minimize bias in our tracking, we applied the *SIFT* algorithm (R. E. Smith et al., 2013) to refine tractograms by selectively discarding streamlines that poorly fit the subject FODs derived from CSD (J.-D. Tournier et al., 2007).

#### 2.4.2 Streamline recognition and segmentation

In order to extract WM fiber bundles (*i.e.*, OR, ILF, IFOF and UF) from the generated whole brain tractogram, we applied BundleSeg (St-Onge et al., 2023), a robust and efficient bundle segmentation approach. Unlike established approaches such as RecoBundles (Garyfallidis et al., 2018), which rely on an approximate intermediate clustering step. BundleSeg uses an exact, exhaustive streamline search for bundle assignments. This approach eliminates the need for clustering and parameter tuning, resulting in significantly improved reproducibility and segmentation quality (St-Onge et al., 2023). To apply BundleSeg, we first registered our subjects’ T1-weighted images to the MNI-152 (ICBM 2009c nonlinear symmetrical) T1-weighted average (Fonov et al., 2011) using ANTs linear registration (<u>antsRegistrationSyNQuick.sh</u>) (Avants et al., 2008). It is important to note that individual subject’s T1-weighted images were rigidly aligned to their respective DWI space prior to registration. Each atlas bundle is then individually aligned to the subject’s tractogram through an iterative process that alternates between searching for the most similar streamlines, using the minimum average direct-flip (MDF) (Garyfallidis et al., 2012) distance and a K-D tree based on the Fast Streamline Search (FSS) framework—and refining alignment with streamline-based registration (Garyfallidis et al., 2015). This search and alignment cycle is repeated, gradually decreasing the search radius at each iteration, until only streamlines within the desired anatomical similarity threshold are retained for each bundle (St-Onge et al., 2023).

#### 2.4.3 DKI-based tractometry and profile characterization

Tract-specific profiling was performed by clustering each extracted bundle using QuickBundles (Garyfallidis et al., 2012) to generate tract centroids. Each centroid represents a cluster of similar streamlines within the bundle, enabling the characterization of tract properties along its length. Each bundle was resampled at 100 equidistant points or nodes (0-99), and DKI metrics were extracted at each node along these streamlines. In addition, the diffusion properties along fiber bundles were summarized using the first four statistical moments: mean, standard deviation (SD), skewness, and kurtosis (Amunts et al., 1999). These metrics help reveal systematic changes in the microstructure of the visual WM tracts (Kasa et al., 2022).

#### 2.4.4 Tractometry–Ophthalmic measure correlations

To assess whether tractometry-derived segmental alterations relate to standard clinical ophthalmic metrics, we computed Pearson correlations between microstructural changes in each bundle segment and retinal nerve fiber layer (RNFL) and ganglion cell layer (GCL) thickness. Specifically, we correlated left-hemisphere bundles (OR, ILF, IFOF) with left- and right-eye RNFL and GCL measurements and repeated the analysis for the right-hemisphere bundles.

### 2.5 Deep MK-Curve corrected DKI tractometry

Since an array of studies have identified glaucoma as a network disorder, we deployed Detect (Chamberland et al., 2021) which is a data-driven unsupervised normative modeling framework based on deep autoencoders (Hinton & Salakhutdinov, 2006), to identify potential relationship between OR and the adjacent WM bundles (*i.e.*, ILF and IFOF) (Herbet et al., 2018; Sarubbo et al., 2013). These bundles were chosen since they are known to be involved in high-order visual processing functions (Herbet et al., 2018; Sarubbo et al., 2013). In addition, considering these bundles as part of the broader visual processing network allows us to explore their interactive relationships, which may collectively help identify anomalies.

Autoencoders, a type of artificial neural network often used for dimensionality reduction, excel at capturing non-linear relationships among input features by learning a compressed, lower-dimensional representation of the data. Their primary function is to generate outputs that closely match the inputs by minimizing reconstruction error (or mean absolute error MAE) (Chamberland et al., 2021). This capability is particularly valuable for anomaly detection, as deviations in the reconstruction can be analyzed. Here the autoencoders were trained on MK-Curve corrected tract profiles from the glaucoma patients and healthy controls (calculated in step 2.4.3). We trained Detect to recognize normative feature sets derived from age/sex matched healthy brain tract profiles (80% training set). After training, the network is tested with an unseen mixture of healthy and patients’ tract profiles (20% validation set) and subsequently exposed to profiles from individuals diagnosed with glaucoma and are on treatment. To derive conservative estimates and assess variations within the model, we repeated this process 100 times and reported the mean MAE for each patient. The performance of the autoencoder was compared to (1) a conventional z-score-distribution approach and (2) principal component analysis (PCA) with Mahalanobis distance, a standard method in cluster analysis and classification. By applying Detect, it also allowed us to identify anomalies at specific locations in multiple tract profiles at subject specific level, which extended beyond traditional group-level analysis in glaucoma.

### 2.6 Statistical analysis

Linear mixed model (LMM) was used to statistically assess group-wise differences in terms of WM profiles for each of the individual MK-Curve corrected DKI parameters. A total of 100 LMMs were run per bundle type (one per node along the length of the tract). Each individual DKI parameter (*e.g.*, MK) was analyzed as the response variable with group, age, and sex as fixed effects, and subject as a random effect. This approach accounted for correlations between data points from the same subject. We used this model to identify differences in the microstructural measures between glaucoma patients and controls in specific bundle segments. We applied False Discovery Rate (FDR) correction to control for multiple comparisons, adjusting for 100 segments per tract with a threshold of *p*FDR<0.05. Output p-values were saved as -log10(*p*) for visualization. The SciPy (v1.11.4) Python package was used to compare tract profiles between patients and controls using the Mann–Whitney U test, after assessing for normality and variance. Finally, the performance of the AutoEncoder, PCA, and z-score methods was evaluated using the mean area under the receiver operating characteristic curve (AUC), which reflects each model’s ability to distinguish between patients and controls across iterations. Group differences in AUC were assessed using two-tailed t-tests.

## 3. Results

### 3.1 MK-Curve correction enhances DKI quality in UKBB dataset

MK-Curve correction led to visible and quantitative improvements in DKI parametric maps. Uncorrected MK images contained anatomically implausible voxels, particularly within regions of complex tissue configuration (*e.g.*, crossing fibers). These voxels were corrected after MK-Curve processing, resulting in more anatomically consistent maps (Fig. 2, top row). Similar improvements were observed in RK and AK. Violin plots comparing MK, RK, and AK values extracted from the left optic radiation revealed that MK-Curve correction reduced extreme outliers and produced tighter, more biologically plausible distributions (Fig. 2, bottom row). Using a repeated-measures linear mixed-effects model (subjects as random intercepts), corrected maps showed significantly higher MK, RK, and AK values than uncorrected data (*p*<0.001). This correction improved the consistency of kurtosis measures across participants, particularly in regions prone to partial-volume effects or fiber crossings. Similar trends were observed for the right optic radiation further demonstrating that MK-Curve correction can be used for microstructural characterization in glaucoma.

**Figure 2.**
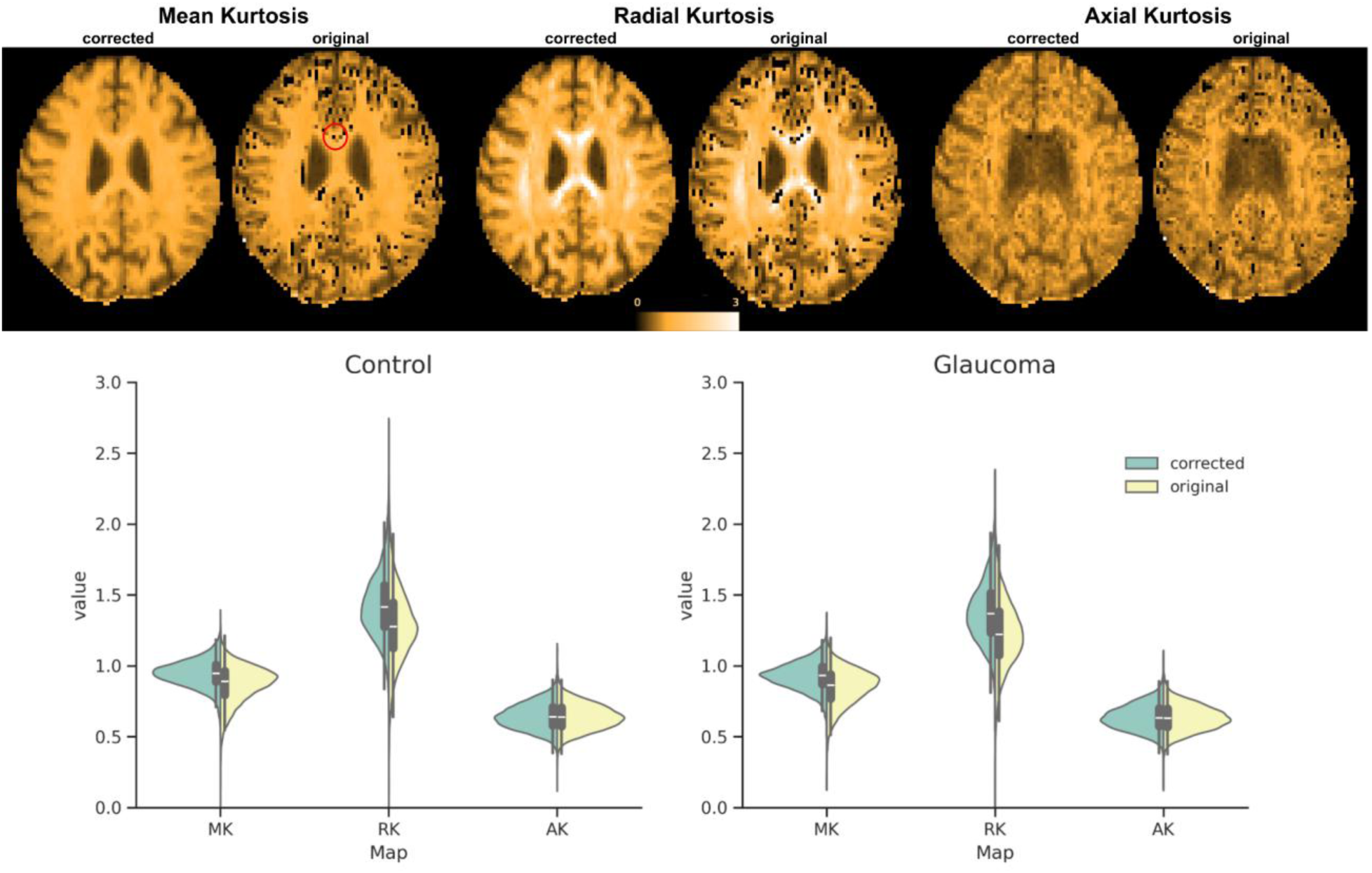
Comparison between MK-Curve corrected and original (uncorrected) DKI maps (mean kurtosis (MK), radial kurtosis (RK) and axial kurtosis (AK)) generated from the UKBB diffusion MRI dataset. The top row shows representative axial slices from a healthy control for corrected and original MK, RK and AK. The red circle on the uncorrected MK slice highlights voxels (dark spots) that were detected and corrected with the MK-Curve technique. The bottom row shows violin plots with error bars comparing corrected (green) vs original MK, RK and AK values extracted from the left optic radiation. Similar results were observed for the right optic radiation. Using linear mixed-effects models with subject as a random effect, corrected maps showed significantly higher MK, RK, and AK values compared to uncorrected data (*p*<0.001), in both the patients group and controls.

### 3.2 MK-Curve corrected DKI-based tractometry and profile characterization

We compared the selected bundles by running LLM for each node (*i.e*., 100 LLM/bundle), starting from point 0 to 99, terminating at the WM-GM interface (*i.e*., to avoid partial volume effect) near the occipital lobe area. Significant group differences in corrected DKI parameters were observed along OR, IFOF and ILF in glaucoma patients compared to healthy controls (Fig.3). Mean kurtosis was significantly higher at multiple nodes along these bundles (Fig. 3A) in comparison to MD (Fig. 3B). The FDR corrected *p*<0.001 are color coded to indicate the locations along the bundle with significant differences and are also mapped on the specific nodes along the bundles for visualization. While only MK and MD profiles are shown here, significant differences were also detected in radial kurtosis (RK) (see Supplementary Fig. 1). As expected, no differences were found in the uncinate fasciculus (UF), which served as a control bundle.

**Figure 3.**
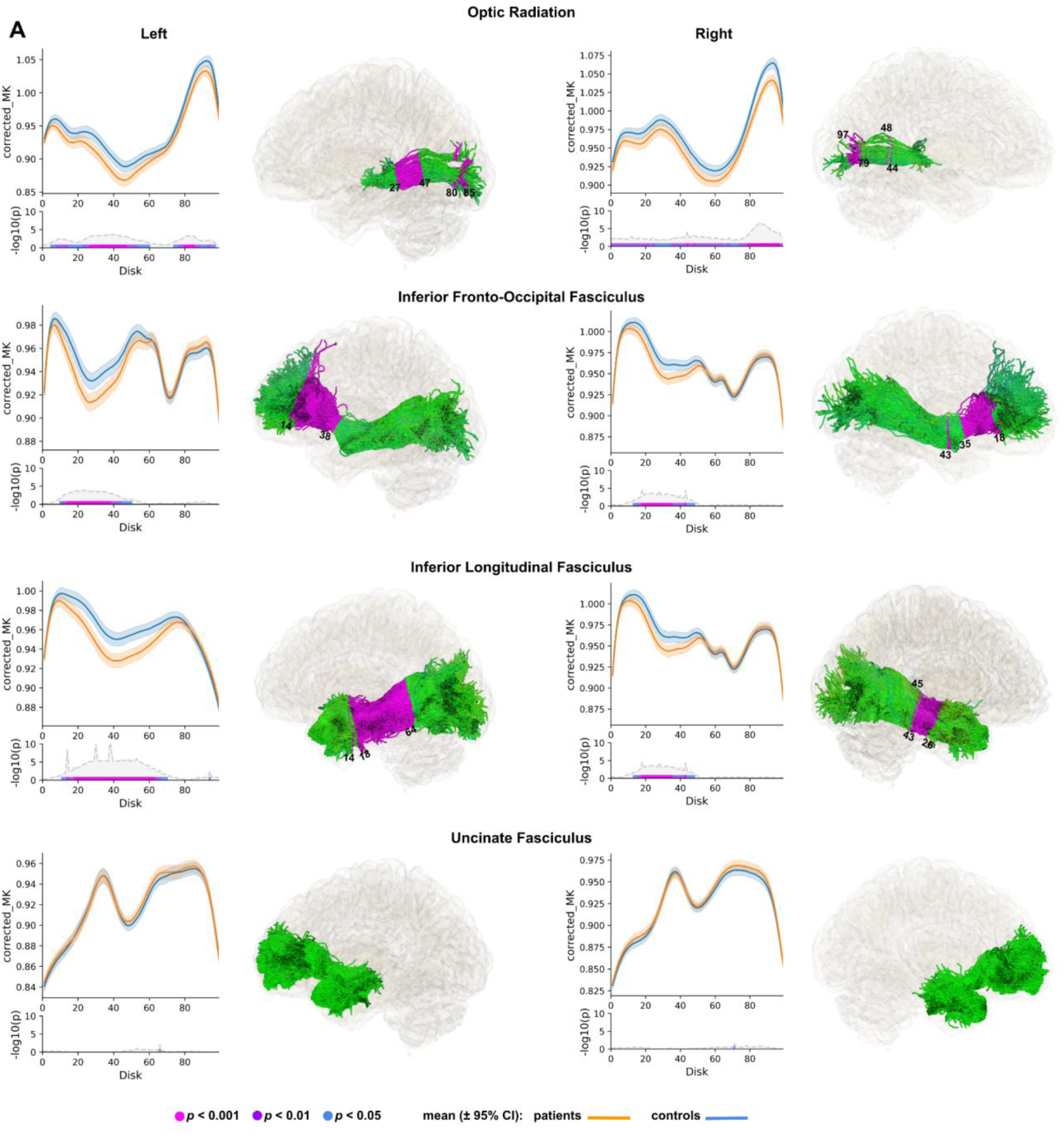
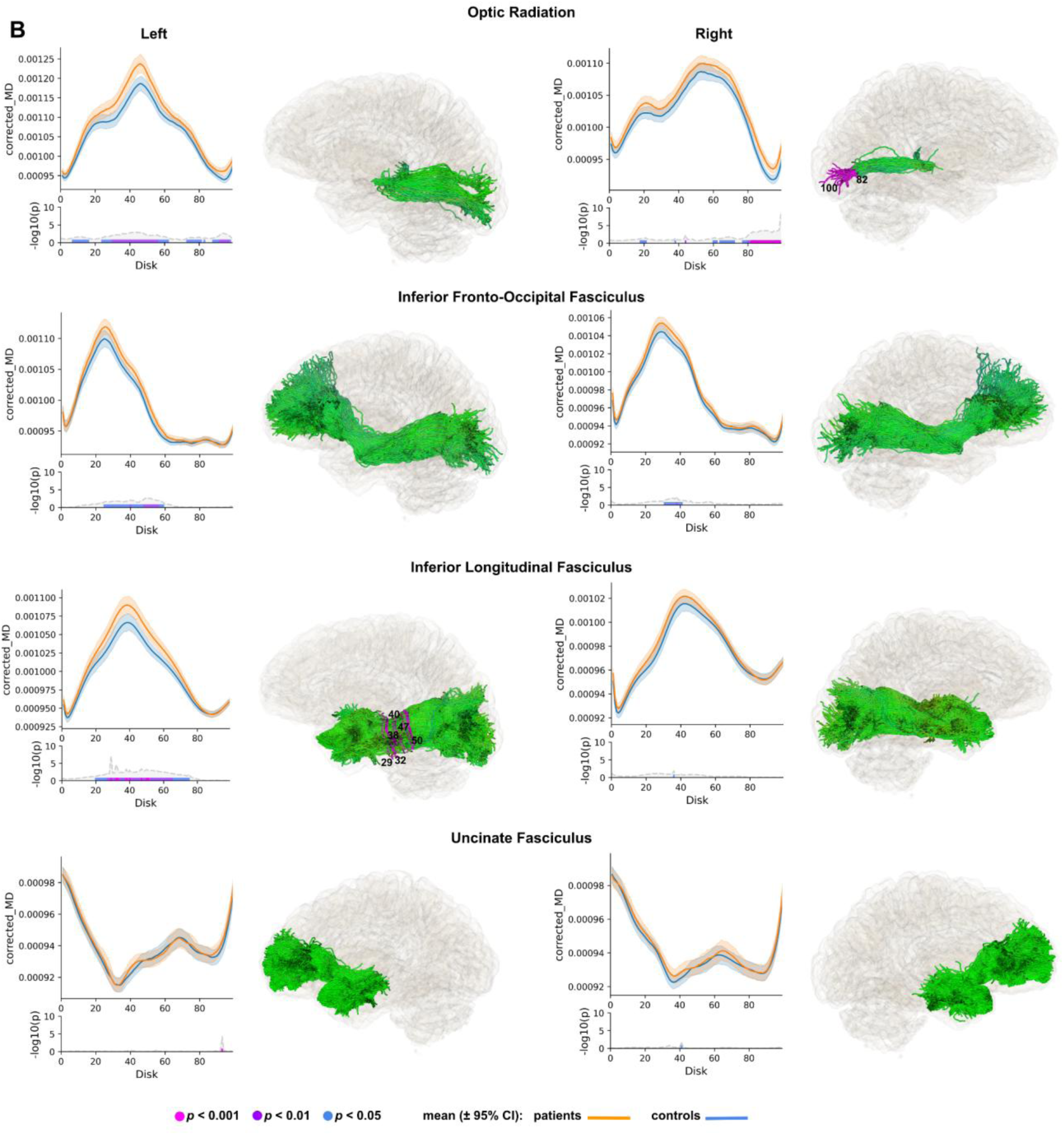
Tractometry analysis reveals localized microstructural abnormalities in glaucoma using MK-Curve corrected DKI parameters. Showing only (A) mean kurtosis (MK) and (B) mean diffusivity (MD). Along-tract profiles of corrected MK and MD are shown for glaucoma patients (orange) and healthy controls (blue) across 100 equidistant nodes along optic radiation (OR) and inferior fronto-occipital fasciculus (IFOF), inferior longitudinal fasciculus (ILF), and the control bundle, uncinate fasciculus (UF). The line and error bars represent the mean and 95% confidence interval, respectively. Node-wise statistical comparisons were performed using linear mixed models tested with FDR correction; –log10(*p*) plots with significance bars (magenta: *p*<0.001; blue: *p*<0.01; black: *p*<0.05) indicate locations of group differences. To visualize the profile differences at corresponding anatomical locations along the bundles (0– 99), the respective p-values (*i.e*., *p*<0.001) are mapped onto the corresponding nodes along the fiber bundles.

For MD tractometry analysis, fewer nodes showed significant increase in MD along OR and left ILF, as shown in Fig.3B.

To illustrate the distribution of the MK-Curve corrected diffusion kurtosis along all diffusion directions (Steven et al., 2014), we show MK and MD as representative metrics of non-Gaussian and Gaussian diffusion, respectively, within the OR. Significant differences between patient and control profiles were observed in both the left and right OR, as reflected in the MK mean_x distribution (Fig.4B-C). Notably, kurtosis and skewness were significantly altered in the left OR among patients (Fig.4B), while the right OR showed a significant difference in standard deviation (Fig.4C). For MD, we found mean_x distribution to be the only significant difference within the left OR (Fig.4D). All reported p-values were Mann–Whitney U (MWU) corrected, with *p*<0.001 indicating high statistical significance.

**Figure 4.**
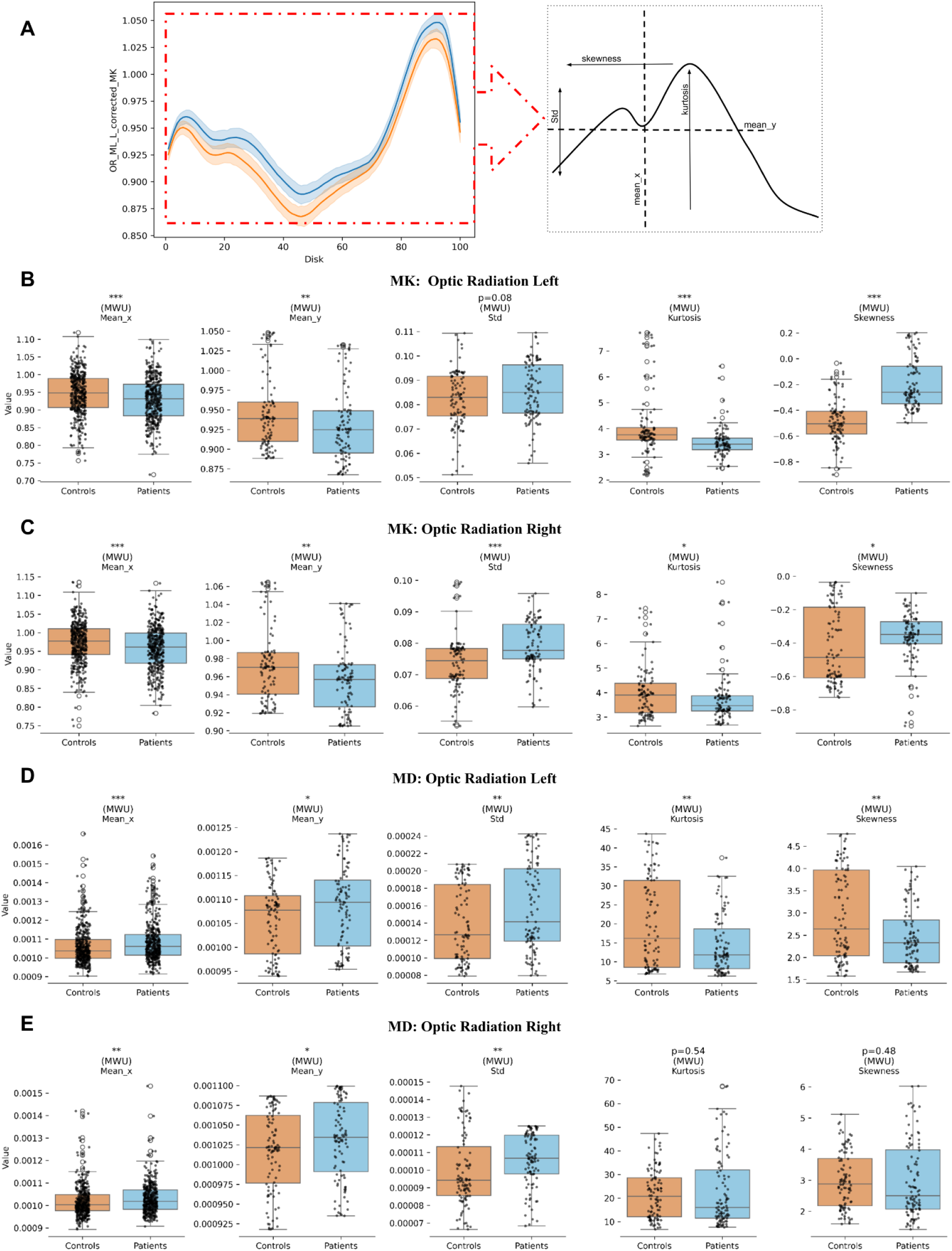
Characterization of DKI-derived quantitative profiles reveals significant differences in mean kurtosis (MK) distribution along the x-direction (mean_x) in both the left (B) and right (C) optic radiation (OR). Panel (A) illustrates the process used to extract tract profile features (see Methods), which are then visualized for MK (B) and mean diffusivity (MD; C) by group –controls (orange) and patients (blue). The x-axes in each subpanel represent the first four statistical moments: mean in the x-direction (mean_x), mean in the y-direction (mean_y), standard deviation (SD), kurtosis, and skewness (rightmost). Only MK and MD metrics are shown, as they exhibited the most pronounced group differences across the OR in both the non-Gaussian (MK) and Gaussian (MD) diffusion components compared to the other bundles. Reported *p*-values were corrected using the Mann–Whitney U (MWU) test; with \*\*\**p* < 0.001 denotes high statistical significance.

### 3.3 Correlation between tractometry and ophthalmic measurements

We next asked whether the node-wise MK alterations in the optic radiation relate to standard optical coherence tomography (OCT) measures of retinal integrity (GCL and RNFL thickness). Across all bundles and both hemispheres, only the right optic radiation’s MK showed a statistically significant but weak relationship with OCT metrics. Specifically, at nodes 48 and 79–83 (the same nodes identified in our group-level tractometry (Fig.5 top)), MK correlated with both GCL and RNFL thickness (Pearson r≈0.10, *p*<0.05) (Fig.5, right panel). No other bundle-metric combinations reached significance. These findings suggest that focal microstructural changes in the right optic radiation partially reflect retinal degeneration, underscoring the potential of combined DKI tract profiling and OCT for multimodal characterization of glaucomatous neurodegeneration.

**Figure 5.**
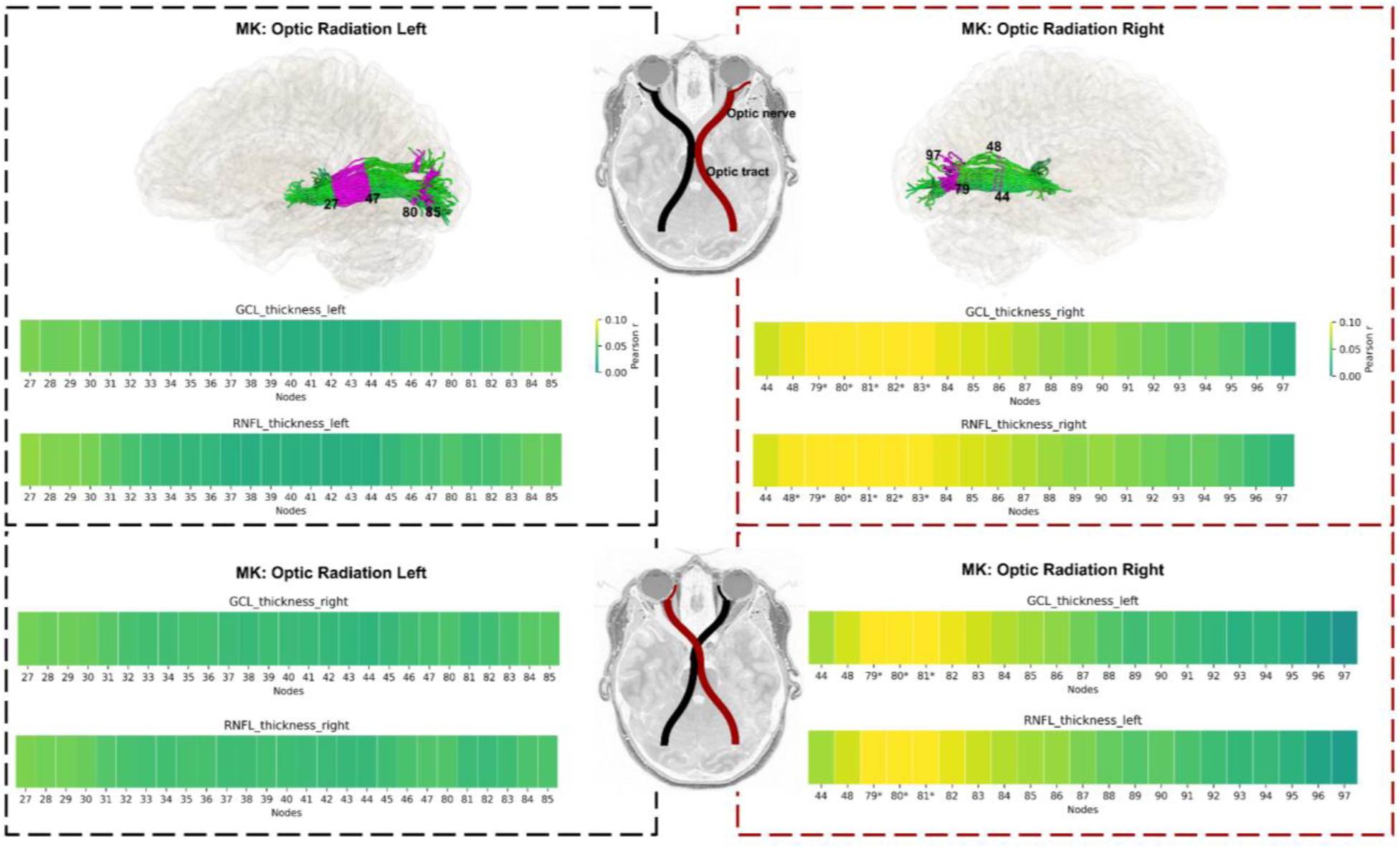
Node-wise correlations between MK in optic radiation (OR) and OCT measures. Heatmaps show Pearson r values for MK at each node (27–85 for left OR, 44–97 for right OR) versus GCL and RNFL thickness. Right-panel: Only the right OR exhibited significant correlations (nodes 48 and 79–83; \**p* < 0.05), with r ≈ 0.10. No other bundle– metric combinations reached significance. The central schematic indicates the anatomical course of the optic nerve - optic tract and then to optic radiation (not shown).

### 3.4 Anomaly detection with Deep MK-Curve corrected DKI tractometry

The Fig.6A schematic illustrates the deep autoencoder (AE) framework we applied for detecting anomalies along the OR and the other non-visual pathway bundles (ILF, IFOF and UF) as implemented in (Chamberland et al., 2021). Firstly, the framework was trained to learn normative sets of features extracted from our healthy subjects’ tract profiles (see section 2.4.3). Once trained, the network is then tested on an unseen mixture of healthy and patients’ tract profiles, and finally applied to profiles from the glaucoma group. Importantly, the model is never exposed to diagnostic labels during training, making this anomaly detection approach entirely unsupervised.

**Figure 6.**
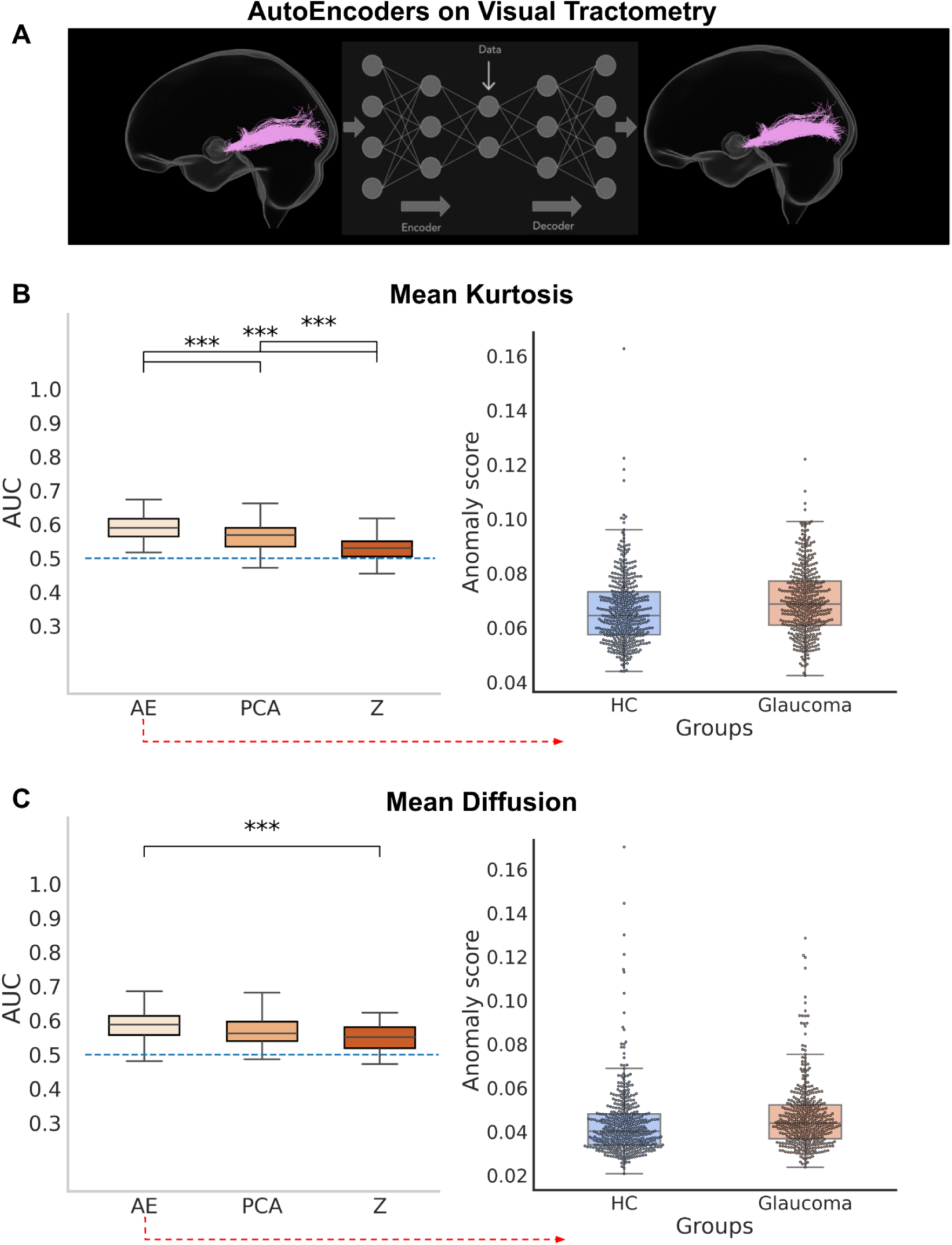
Deep autoencoder-based anomaly detection applied to tractometry data. (A) Schematic of the autoencoder (AE) framework trained on healthy tract profiles to learn normative diffusion patterns across the optic radiation (OR), inferior longitudinal fasciculus (ILF), inferior fronto-occipital fasciculus (IFOF), and uncinate fasciculus (UF). The model is fully unsupervised and never exposed to diagnostic labels. (B–C) Discrimination performance (left) and anomaly scores (right) for mean kurtosis (MK, B) and mean diffusivity (MD, C). AE outperformed PCA and z-score approaches in distinguishing glaucoma from healthy controls based on AUC scores (\*\*\**p* < 0.001, Bonferroni corrected). MK showed the strongest separation in anomaly scores between groups.

To assess the discriminative power of the autoencoder for investigating glaucoma patients white matter microstructure we used the AUC scores across the DKI metrics. The autoencoder approach showed higher sensitivity-specificity after 100 iterations than PCA with Mahalanobis distance and z-score. For example, the MK features showed higher discriminating power for autoencoder (AUC=0.61, \*\*\**p*<0.001, Bonferroni corrected with α = 0.01, two-tailed t-tests for AE–PCA and AE–z, respectively) compared to conventional approaches, PCA with Mahalanobis distance (AUC=0.57) and z-score (AUC=0.54) as shown in the boxplots in Fig.6(B-left). The MK features show higher reconstruction error for the glaucoma group (Fig.6 (B-right)) compared to healthy controls (orange boxplot). Autoencoder trained with MD features again outperforms PCA and z-score methods (AUC=0.59, \*\*\**p*<0.001 for AE–PCA and AE–z, respectively), though the anomaly score separation between groups is less pronounced than for MK (Fig.6C). These findings demonstrate the enhanced sensitivity of deep autoencoders compared with traditional linear univariate approaches (*i.e*., PCA and z-score) in detecting tract-level microstructural changes, with MK yielding greater discriminative power than MD.

A key advantage of using an autoencoder in this study is its ability to detect subject-level white matter anomalies—complementing our group-level findings. To demonstrate this capability, we selected a glaucoma patient with a high polygenic risk score (PRS), given that elevated PRS is associated with increased susceptibility to primary open-angle glaucoma (POAG) (Craig et al., 2020; Singh et al., 2024). Figure 7A illustrates the PRS distribution, highlighting the selected patient within the top 5% of genetic risk.

**Figure 7.**
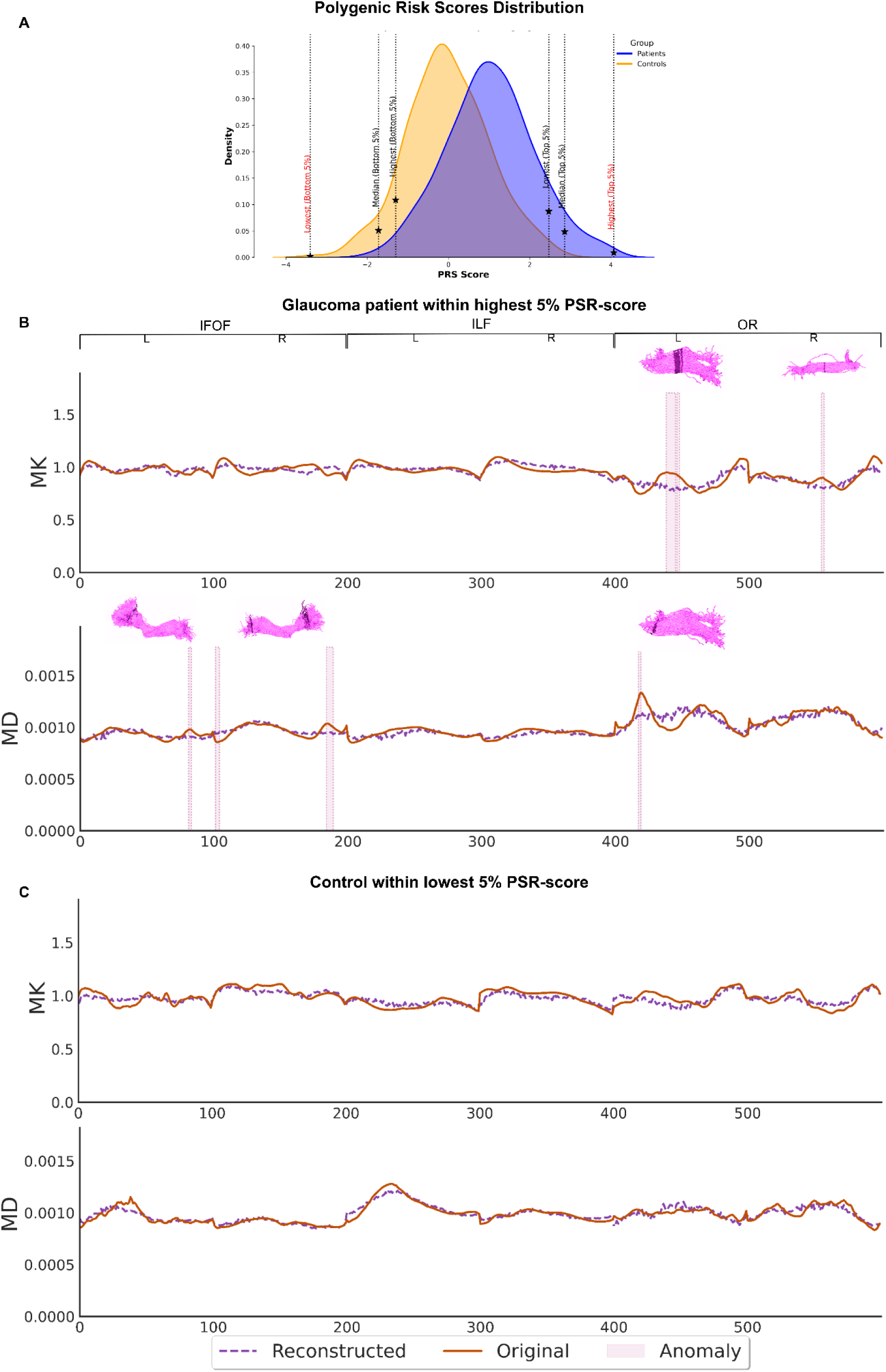
Subject-level anomaly detection based on polygenic risk for glaucoma. (A) Distribution of polygenic risk scores (PRS) for glaucoma patients and controls (orange), as reported in the UKBB dataset (Sudlow et al., 2015). A patient within the top 5% of PRS values was selected for individual analysis (red font and black dashed line with *). (B) Mean kurtosis (MK, top) and mean diffusivity (MD, bottom) profiles for the selected patient across the inferior fronto-occipital fasciculus (IFOF), inferior longitudinal fasciculus (ILF), and optic radiation (OR). Original profiles (orange) are overlaid with autoencoder reconstructions (purple dashed). Anomalies (pink shaded regions) were detected using leave-one-out cross-validation, reporting nodes with at least two consecutive outliers. Detected anomalies were mapped back onto fiber tracts (indicated as dark purple), showing that deviations aligned with regions identified in group-level analyses (see Fig. 3), highlighting the utility of this subject-specific framework. (C) MK (top) and MD (bottom) profiles for a control subject in the lowest 5% of PRS (red font and black dashed line with *). No anomalies were detected across the IFOF, ILF, or OR, reinforcing the method’s specificity in distinguishing between high- and low-risk individuals.

We then compared this individual against the rest of the cohort, identifying localized anomalies in MK (Fig.7B-top) and MD (Fig.7B-bottom) features across the IFOF, ILF, and OR. Anomalous nodes were detected using a leave-one-out cross-validation approach, with results reported for regions showing at least two consecutive outliers. These anomalies are highlighted in pink along the node-wise representation of each bundle (nodes 0–99). MK anomalies were most prominent in the left OR, with additional subtle deviations observed in the right OR, including MD features (Fig. 7B, bottom). To spatially contextualize these findings, the anomalies were mapped back onto the fiber tracts of the selected subject. Notably, the locations of these deviations aligned with regions identified in our group-level tractometry analysis (Fig. 3), demonstrating the added value of autoencoder-based anomaly detection for individual assessment beyond traditional group comparisons.

For further comparison, we also evaluated a control subject within the lowest 5% of the PRS distribution (Fig. 7A). No anomalies were detected across any of the examined tracts in this individual (Fig. 7C), supporting the specificity of the autoencoder in identifying microstructural abnormalities associated with increased genetic risk for glaucoma.

## 4 Discussion

This study introduces a novel framework that combines MK-Curve-corrected diffusion kurtosis imaging, tractometry, and deep autoencoder-based normative modeling to detect microstructural white matter anomalies in glaucoma. By integrating advanced diffusion modeling with individualized anomaly detection, our approach enables both sensitive group-level comparisons and subject-specific assessments of disease-related changes along white matter tracts. These findings provide new insights into the spatially distributed nature of glaucomatous neurodegeneration and highlight the clinical potential of personalized tract profiling in neuro-ophthalmology –validating our hypothesis and extending previous group-level findings (Kruper et al., 2024; Nucci et al., 2020; Z. Sun et al., 2020).

### 4.1 Enhancing DKI sensitivity with MK-Curve correction in UKBB

Our results demonstrate that MK-Curve correction enhances the anatomical plausibility and reliability of DKI metrics in the UKBB dataset. Uncorrected MK maps frequently showed implausibly low kurtosis values in complex tissue organization, particularly in regions with crossing or fanning fibers and deep white matter – issues previously noted in diffusion studies (Henriques et al., 2021; Tabesh et al., 2011). MK-Curve correction effectively removed these artifacts, yielding more consistent MK, RK, and AK maps. This was reflected in the reduced outlier presence and tighter distribution of values, particularly within the optic radiation. These improvements are critical in large-scale diffusion studies, where minimizing artifact-driven variability strengthens the interpretability of group comparisons and downstream modeling (Ades-Aron et al., 2025; Henriques et al., 2021). Our findings support MK-Curve as a valuable preprocessing step for tract-based DKI analysis in glaucoma.

### 4.2 Improved DKI Tractometry via MK-Curve Correction

MK-Curve corrected DKI revealed localized microstructural abnormalities in glaucoma patients along the OR, IFOF, and ILF bundles that are critical for visual and visual cognitive processing (Herbet et al., 2018; Rokem et al., 2017; Sarubbo et al., 2013). By applying linear mixed models (LMMs) at 100 equidistant nodes per tract, we found that MK was significantly decreased at multiple nodes in these bundles in glaucoma patients, compared to healthy controls. These spatially detected abnormalities were not observed in our control bundle, UF, thereby reinforcing the disease specificity of our findings.

The heightened sensitivity of MK relative to MD aligns with prior evidence suggesting that DKI-derived metrics better capture the non-Gaussian diffusion patterns characteristic of neurodegeneration (Fieremans et al., 2011; Steven et al., 2014; Z. Sun et al., 2020). MK increases may reflect glial hypertrophy or extracellular matrix remodeling in response to transsynaptic degeneration originating from retinal ganglion cell loss—an established hallmark of glaucoma (Gupta & Yücel, 2007; Si et al., 2025). The presence of these microstructural changes within both primary and associative visual tracts reinforces the hypothesis that glaucoma is a brain-wide disorder rather than an eye-specific disease (Frezzotti et al., 2014; Nucci et al., 2013).

Importantly, the UF, which is not directly implicated in visual processing, showed no significant group differences. This supports the regional specificity of our findings and highlights the clinical potential of DKI-based tract profiling in glaucoma. Furthermore, the inclusion of RK in our supplementary analysis (Supplementary Fig. 1) showed complementary group differences, particularly in regions of complex fiber architecture, further supporting the utility of DKI for probing subtle changes not captured by conventional DTI metrics. These findings underscore the potential of DKI tractometry as a non-invasive biomarker to detect early white matter changes in glaucoma, potentially prior to overt visual field loss. Mapping localized microstructural changes along anatomically defined tracts offers a promising avenue for monitoring and evaluating the efficacy of neuroprotective interventions.

To further characterize microstructural changes along the OR, we analyzed the full distribution of MK and MD using the first four statistical moments. Glaucoma patients showed significant alterations in MK distributions: increased mean, kurtosis, and skewness in the left OR, and elevated standard deviation in the right OR (*p*<0.001, Bonferroni-corrected). These results highlight MK’s sensitivity to spatial heterogeneity in WM and emphasize the value of distributional profiling over traditional mean-based comparisons. In contrast, MD differences were limited to a modest mean increase in the left OR, reinforcing MK’s superior sensitivity to the microstructure.

### 4.3 Correlation between tractometry and ophthalmic measurements

The observed weak but significant correlations between MK in the right optic radiation and OCT measures of retinal integrity suggest that microstructural alterations captured by MK-Curve– corrected DKI partially reflect the degree of retinal ganglion cell and nerve fiber layer loss in glaucoma, supporting previous ROI based findings (Z. Sun et al., 2020). Specifically, only the right OR exhibited node-wise MK associations with both GCL and RNFL thickness (r ≈ 0.10, *p*<0.05 at nodes 48 and 79–83), corresponding precisely to the nodes of significant group-level tractometry changes. While the effect sizes are small, these findings provide preliminary evidence that focal diffusion kurtosis changes along the OR are linked to established clinical biomarkers of glaucomatous neurodegeneration. The modest correlation strength underscores the multifactorial nature of OR pathology, where DKI metrics may capture complementary aspects of white matter integrity not fully accounted for by retinal measures alone. Nonetheless, integrating tract-specific DKI profiling with OCT holds promise for a multimodal approach to characterizing the full extent of neurodegenerative changes in glaucoma.

### 4.4 Anomaly detection with Deep MK-Curve corrected tractometry

Our results demonstrate that deep autoencoder-based normative modeling, when applied to MK-Curve-corrected DKI tractometry, provides enhanced sensitivity for detecting microstructural deviations in glaucoma. Compared to conventional linear approaches (*e.g.*, PCA with Mahalanobis distance and z-score), the autoencoder yielded significantly higher AUC scores, particularly for MK, underscoring its superior discriminative power for non-Gaussian diffusion changes, consistent with previous studies emphasizing the diagnostic value of higher-order diffusion metrics (Fieremans et al., 2011; Steven et al., 2014; Gong et al., 2013). The unsupervised design of the model allows for anomaly detection without diagnostic labels, offering a robust framework for individualized assessment. Subject-level application in a genetically high-risk glaucoma patient further revealed focal anomalies in MK and MD, especially along the optic radiation and associative visual pathways, which spatially overlapped with group-level tractometry findings. This convergence highlights the clinical potential of deep learning approaches to extend beyond traditional group-based analyses and toward precision mapping of white matter pathology (Chamberland et al., 2021; Pinaya et al., 2019; Huizinga et al., 2018). Given the progressive nature of glaucoma and its early CNS involvement, subject-level anomaly detection may facilitate early diagnosis and tracking of disease progression, particularly in genetically at-risk populations (Gupta and Yücel, 2007; Singh et al., 2024). Our findings support the integration of advanced modeling techniques into diffusion MRI pipelines for individualized neuroimaging biomarkers in neuro-ophthalmology and beyond.

### 4.5 Limitations and future directions

While our findings highlight the potential of MK-Curve-corrected DKI tractometry and deep learning-based anomaly detection for identifying white matter alterations in glaucoma, several limitations warrant consideration. First, although the UKBB dataset offers high-quality diffusion MRI and a large sample size, its cross-sectional nature limits causal inference. Longitudinal studies are necessary to determine whether the observed microstructural changes precede or follow clinical manifestations of glaucoma. Additionally, the lack of more precise glaucoma diagnosis in UKBB also led to assuming changes to visual tracts were mirrored across both the left and right eye (bilateral disease), when many individuals with less severe disease might have only unilateral visual complications. Second, while the MK-Curve correction improves DKI reliability, residual artifacts in regions with extreme partial volume effects or high noise may still influence metric estimation. Third, our autoencoder framework, although unsupervised and generalizable, was only applied to a limited set of bundles and diffusion metrics. Including additional white matter tracts and multi-parametric map features could improve sensitivity and coverage. Finally, the clinical applicability of subject-level anomaly detection needs further validation in independent cohorts and in conjunction with functional or behavioral outcomes. Future studies integrating multimodal imaging, visual field data, and genetic risk profiling will be important for translating these methods into personalized diagnostic tools.

## 5. Conclusions

This study presents a novel framework combining MK-Curve-corrected diffusion kurtosis imaging, tractometry, and deep autoencoder-based normative modeling to detect localized white matter abnormalities in glaucoma. We demonstrate that MK-Curve correction enhances DKI reliability, and that MK offers greater sensitivity than MD in identifying disease-related changes along visual pathways. Autoencoder-based anomaly detection further enables subject-specific assessment, outperforming traditional methods and aligning well with group-level findings. These results support the potential clinical feasibility of this approach for early detection, monitoring, and personalized assessment of neurodegeneration in glaucoma.

## Data Availability

All data produced in the present study are available upon reasonable request to the authors

## Data and Code Availability

The data used in this study were obtained from the UK Biobank (UKBB) resource (Sudlow et al., 2015) accessible at https://www.ukbiobank.ac.uk/enable-your-research/about-our-data.

The code for the anatomical guided tracking and analysis is implemented as a Snakemake pipeline at: https://github.com/Loxlan/glaucoma_smk.git

## Author Contributions

Conceptualization: L.W.K., W.S., S.H., E.K. and H.V.D.M. Methodology: L.W.K Software: L.W.K Validation: L.W.K Formal analysis: L.W.K Investigation: L.W.K Writing—Original Draft: L.W.K Writing—Review & Editing: L.W.K. W.S. S.H., E.K. and H.V.D.M. Visualization: L.W.K Data Curation: L.W.K and W.S Project administration: L.W.K and E.K Funding acquisition: S.H. and H.V.D.M.

## Funding

This work was supported by Vision Research Foundation, New Zealand and Rapanui Trust, Tairāwhiti Gisborne, New Zealand.

## Declaration of Competing Interest

The authors report no competing interests.

## Acknowledgements

L.W.K. was supported by a Research Fellowship and W.S. by a Senior Research Fellowship, both awarded by the Vision Research Foundation. E.K was supported by the Hugh and Moira Green Senior Research Fellowship

The author(s) wish to acknowledge the use of New Zealand eScience Infrastructure (NeSI) high performance computing facilities, https://www.nesi.org.nz, and the support of Kānoa - Regional Development Unit of Aotearoa New Zealand

## Supplementary Data

**Supplementary Figure 1.**
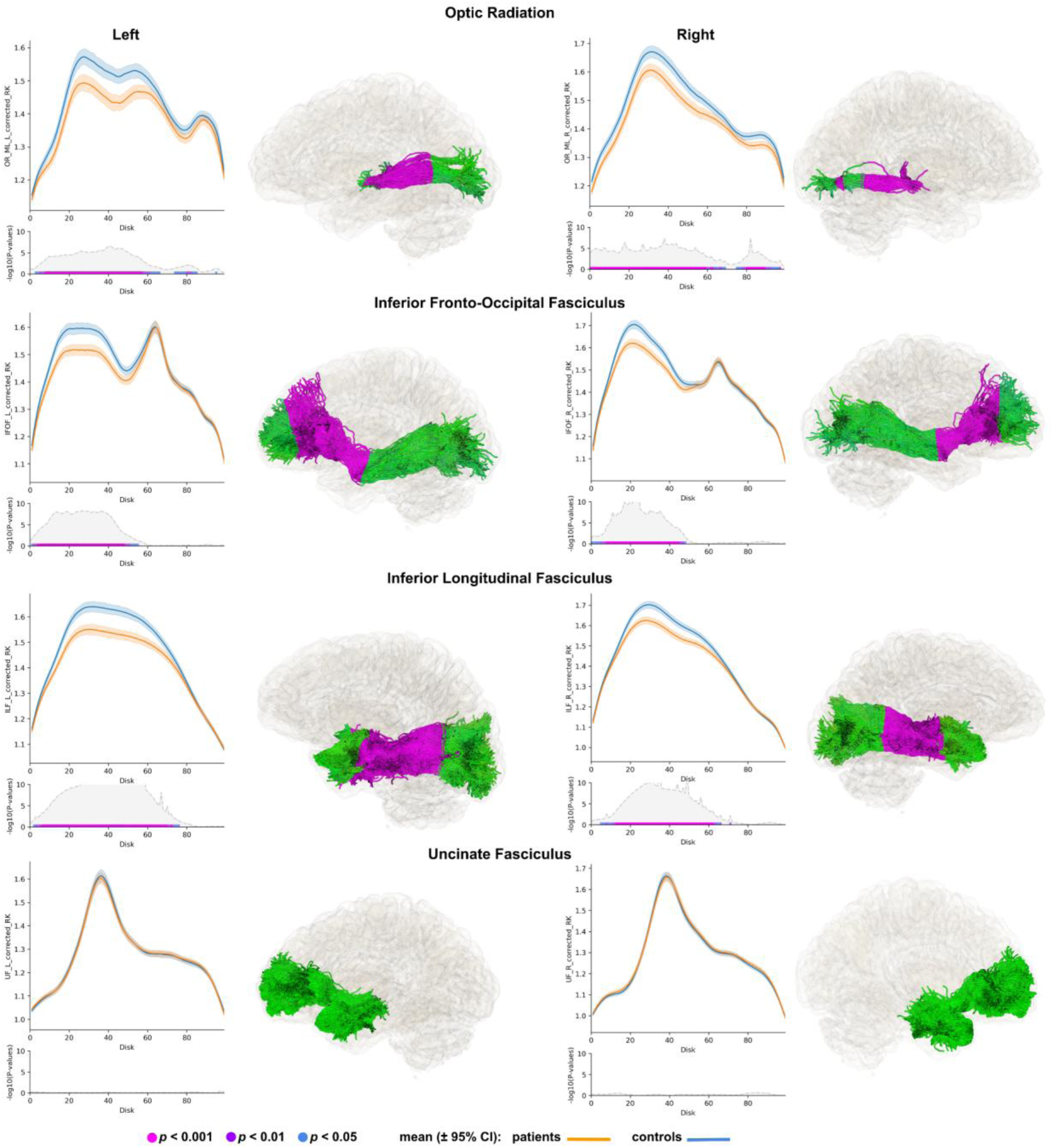
Tractometry analysis reveals localized microstructural abnormalities in glaucoma using MK-Curve-corrected DKI parameters. Along-tract profiles of corrected RK shown for glaucoma patients (orange) and healthy controls (blue) across 100 equidistant nodes along optic radiation (OR) and inferior fronto-occipital fasciculus (IFOF), inferior longitudinal fasciculus (ILF), and the control bundle, uncinate fasciculus (UF). The line and error bars represent the mean and 95% confidence interval, respectively. Node-wise statistical comparisons were performed using linear mixed models tested with FDR correction; –log10(p) plots with significance bars (magenta: *p*<0.001; blue: *p*<0.01; black: *p*<0.05) indicate locations of group differences. To visualize the profile differences at corresponding anatomical locations along the bundles (0–99), the respective p-values are mapped onto the corresponding nodes along the fiber bundles.

## Notes

### Competing Interest Statement

The authors have declared no competing interest.

### Funding Statement

This study was funded by The Vision Research Foundation, Auckland, New Zealand and Rapanui Trust, Gisborne, New Zealand

### Author Declarations

The source dataset for this study was downloaded from UK Biobank a public database. This dataset existed before the study was formulated. Here is the link to the database and information on how to download or use the dataset https://www.ukbiobank.ac.uk/enable-your-research/about-our-data

